# COVID-19 ARDS is characterized by a dysregulated host response that differs from cytokine storm and may be modified by dexamethasone

**DOI:** 10.1101/2020.12.28.20248552

**Authors:** Aartik Sarma, Stephanie A. Christenson, Eran Mick, Catherine DeVoe, Thomas Deiss, Angela Oliveira Pisco, Rajani Ghale, Alejandra Jauregui, Ashley Byrne, Farzad Moazed, Natasha Spottiswoode, Pratik Sinha, Beth Shoshana Zha, Paula Hayakawa Serpa, K. Mark Ansel, Jennifer G. Wilson, Aleksandra Leligdowicz, Emily R. Siegel, Marina Sirota, Joseph L. DeRisi, Michael A. Matthay, COMET Consortium, Carolyn M. Hendrickson, Kirsten N. Kangelaris, Matthew F. Krummel, Prescott G. Woodruff, David J. Erle, Carolyn S. Calfee, Charles R. Langelier

## Abstract

We performed comparative lower respiratory tract transcriptional profiling of 52 critically ill patients with ARDS from COVID-19 or other etiologies, or without ARDS. We found no evidence of cytokine storm but instead observed complex host response dysregulation driven by genes with non-canonical roles in inflammation and immunity that were predicted to be modulated by dexamethasone. Compared to other viral ARDS, COVID-19 was characterized by impaired interferon-stimulated gene expression.

## Brief Communication

In its most severe form, coronavirus disease 2019 (COVID-19) can precipitate the acute respiratory distress syndrome (ARDS), which is characterized by low arterial oxygen concentrations, alveolar injury and a dysregulated inflammatory response in the lungs^1^. Early reports hypothesized that COVID-19 ARDS was driven by a “cytokine storm” based on the detection of higher circulating inflammatory cytokine levels in critically ill COVID-19 patients compared to those with mild disease or healthy controls^2^. Recent studies, however, have found that patients with COVID-19 ARDS in fact have lower plasma cytokine levels compared to those with ARDS due to other causes^3^, highlighting a need to understand the underlying mechanisms of COVID-19 ARDS.

Clinical trials have demonstrated a significant mortality benefit for dexamethasone in COVID-19 patients with ARDS^4^, implicating a role for dysregulated inflammation in COVID-19 pathophysiology given the immunomodulatory effects of corticosteroids. In contrast, clinical trials of corticosteroids for ARDS prior to the SARS-CoV-2 pandemic have had mixed results, ranging from benefit to possible harm^1^. These differences suggest distinct biology in COVID-19 ARDS, with important implications for pathogenesis and treatment.

While several studies have assessed host airway transcriptional responses to SARS-CoV-2^5,6^, none have compared COVID-19 ARDS to other causes of ARDS. We hypothesized that this comparison would identify distinct biological features of SARS-CoV-2 related lung injury, and thus conducted a case-control study of 52 critically ill patients requiring mechanical ventilation (**Supplementary Table 1**) for ARDS from COVID-19 (COVID-ARDS, n= 15), ARDS from other etiologies (Other-ARDS, n= 32), or for airway protection in the absence of pulmonary disease (No-ARDS, n = 5). Other ARDS etiologies included pneumonia, aspiration, sepsis, and transfusion reaction.

Patients were enrolled at two tertiary care hospitals in San Francisco, California under research protocols approved by the University of California San Francisco Institutional Review Board (**Supplementary Methods**). We excluded immunosuppressed patients to avoid confounding the measurement of host inflammatory responses (**Supplementary Methods**). Tracheal aspirate (TA) was collected within five days of intubation and underwent RNA sequencing (**Supplementary Methods**).

We compared TA gene expression between COVID-ARDS and Other-ARDS patients (**Supplementary Methods, Figure 1a, Supplementary Data 2)** and identified 696 differentially expressed genes at an adjusted P-value < 0.1, as well as differentially activated pathways using Ingenuity Pathway Analysis (IPA)^7^. Notably, we did not observe elevated expression of genes encoding canonical proinflammatory cytokines, such as IL-1 or IL-6, in COVID-ARDS compared to Other-ARDS. In fact, the IL-1, IL-6 and IL-8 signaling pathways were more highly activated in Other-ARDS, whereas COVID-ARDS patients had comparable inflammatory pathway activation to No-ARDS controls (**Figure 1b, Supplementary Data 3)**. We also found attenuation of the proinflammatory HIF-1a and nitric oxide signaling pathways in COVID-ARDS compared to Other-ARDS patients. To relate these lower respiratory tract findings to systemic inflammatory responses, we also assessed circulating plasma cytokines. We found no difference in IL-6 or IL-8 levels, and lower concentrations of sTNFR1, in COVID-ARDS versus Other-ARDS patients (**Figure 1c, Supplementary Data 4**).

**Figure 1.**
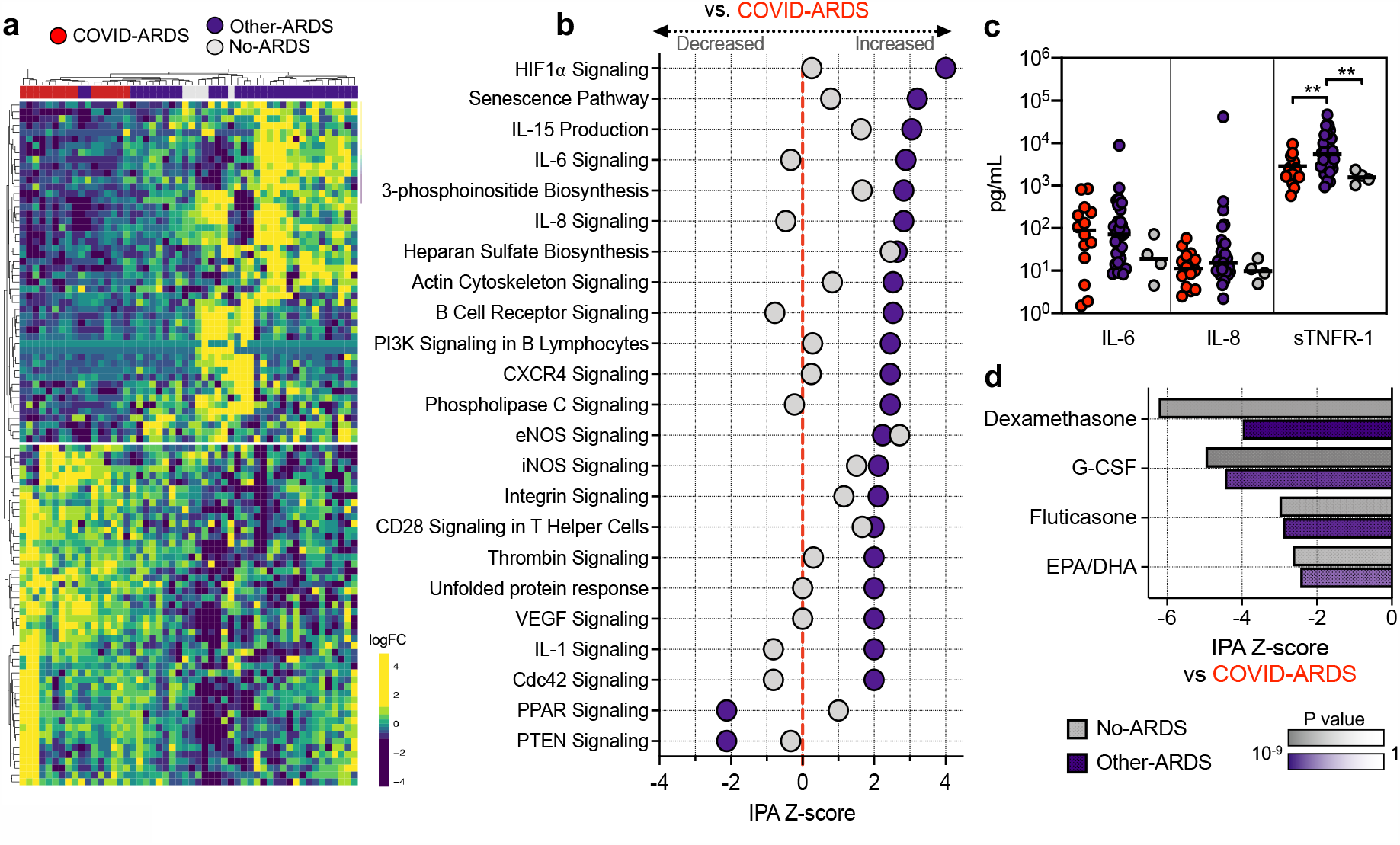
Lower respiratory tract transcriptional signature of COVID-19 ARDS. **a)** Heatmap of the top 50 differentially expressed genes by adjusted p value between patients with COVID-19 related ARDS (COVID-ARDS, red) versus controls with ARDS due to other etiologies (Other-ARDS, violet). Intubated controls with no ARDS were also included in the unsupervised clustering (No-ARDS, grey). **b)** Ingenuity Pathway Analysis (IPA) based on differential gene expression analyses demonstrating expression of signaling pathways by IPA activation Z-score with respect to a baseline of COVID-ARDS. Values are tabulated in (Supplementary Data 3). **c)** Differences in plasma inflammatory cytokine concentrations between patients with ARDS due to COVID-19 (COVID-ARDS, red) or other etiologies (Other-ARDS, violet). Lines indicate medians. P values calculated based on Mann-Whitney test. Values tabulated in (Supplementary Data 4). **d)** Pharmacologic agents predicted to mitigate the dysregulated host response of COVID-19 ARDS with respect to Other-ARDS (violet) or No-ARDS patients (grey) identified by computational matching against the IPA database of drug transcriptional signatures. Values tabulated in (Supplementary Data 5). Pathways with a Z-score absolute value > 2 and overlap P value < 0.05 are significant.

Evaluation of genes with the most significant expression differences in COVID-ARDS compared to Other-ARDS did, however, reveal several differences in genes regulating immunity and inflammation (**Supplementary Data 2**). For instance, among genes upregulated in COVID-ARDS, *P2RY14* functions in purinergic receptor signaling to mediate inflammatory responses and its ligand UDP-glucose promotes neutrophil recruitment in the lung^8^. Conversely, *ARG1*, which promotes macrophage efferocytosis and inflammation resolution^9^, was downregulated in COVID-ARDS versus Other-ARDS patients.

At the pathway level, we observed activation of PTEN signaling in COVID-ARDS compared to both Other-ARDS and No-ARDS patients (**Figure 1B, Supplementary Data 3**). PTEN modulates both innate and adaptive immune responses by opposing the activity of PI3K^10^. Consistent with our observations, PTEN attenuates expression of certain cytokines while amplifying other innate immune responses in a manner that may promote injurious inflammation during respiratory infections^11^, suggesting a potentially pathologic role in COVID-ARDS. *In silico* prediction of cell type composition (**Methods, Supplementary Figure 1, Supplementary Data 5**) did not identify differences in lymphocyte, macrophage or neutrophil populations but did identify markedly decreased proportions of type-2 alveolar epithelial cells and increased proportions of goblet and ciliated cells in COVID-ARDS compared to Other-ARDS. This may reflect alveolar epithelial injury, airway remodeling, and/or preferential SARS-CoV-2 infection of cells with the highest expression of *ACE2* and *TMPRSS2*^12^.

To test the hypothesis that existing pharmaceuticals might counter the dysregulated transcriptional signature of COVID-19 related ARDS, we compared differentially expressed genes against the IPA database of 12,981 drug treatment-induced transcriptional signatures derived from human studies and cell culture experiments^7^ (**Supplementary Methods, Figure 1c, Supplementary Data 6**). Dexamethasone was the compound predicted to most significantly counter-regulate the genes expressed in COVID-ARDS patients compared to No-ARDS patients, and was also significant with respect to the Other-ARDS group. This finding was striking given that dexamethasone is the only drug thus far proven to confer a mortality benefit in patients with severe COVID-19^4^. Granulocyte colony stimulating factor (G-CSF) was also significant, which is intriguing given that a recent clinical trial found a mortality benefit in COVID-19 patients treated with this agent^13^. Other corticosteroids (fluticasone, prednisolone) as well as omega-3 fatty acids (eicosapentenoic and docosahexaenoic acids) were also predicted to counteract the transcriptional profile of COVID-ARDS with respect to comparator groups and thus may represent possible therapeutic agents (**Figure 1c, Supplementary Data 6**).

As our analysis did not reveal evidence of a cytokine storm in COVID-19 ARDS, we hypothesized that other steroid-responsive pathways may be responsible for the therapeutic benefit of dexamethasone. Although commonly thought of as indiscriminate immunosuppressive medications, glucocorticoids affect diverse biological processes. We therefore proceeded to examine the genes comprising the transcriptional signature of COVID-ARDS that were also predicted to be regulated by dexamethasone (**Supplementary Data 7**). Interestingly, both dexamethasone and G-CSF were predicted to attenuate the expression of several genes highly upregulated in COVID-ARDS versus controls (e.g., *P2YR14*) as well as other genes with well-established roles in immunity, inflammation, and interferon responses. For instance, we found that COVID-ARDS patients had increased expression of the interferon-inducible and dexamethasone-regulated gene *EPSTI1*, which promotes M1 macrophage polarization^14^, and *STAT1*, which induces chemokine expression, regulates differentiation of hematopoietic cells, and promotes reactive oxygen species production^15^.

ARDS is a heterogeneous syndrome caused by diverse infectious and non-infectious insults^1^. To more precisely understand host response relationships between subtypes of ARDS, we performed secondary analyses comparing the transcriptional signature of COVID-ARDS without co-infections (n = 8) to that of ARDS caused exclusively by other viral (n = 4, **Figure 2a, Supplementary Data 1**) or bacterial (n = 9, **Figure 2b**) lower respiratory tract infections (LRTI) (**Supplementary Data 8**). COVID-ARDS was characterized by lower expression of proinflammatory signaling pathways compared to bacterial LRTI-associated ARDS **(Figure 2c, Supplementary Data 9)**, but higher levels of the same pathways compared to viral LRTI-associated ARDS.

**Figure 2.**
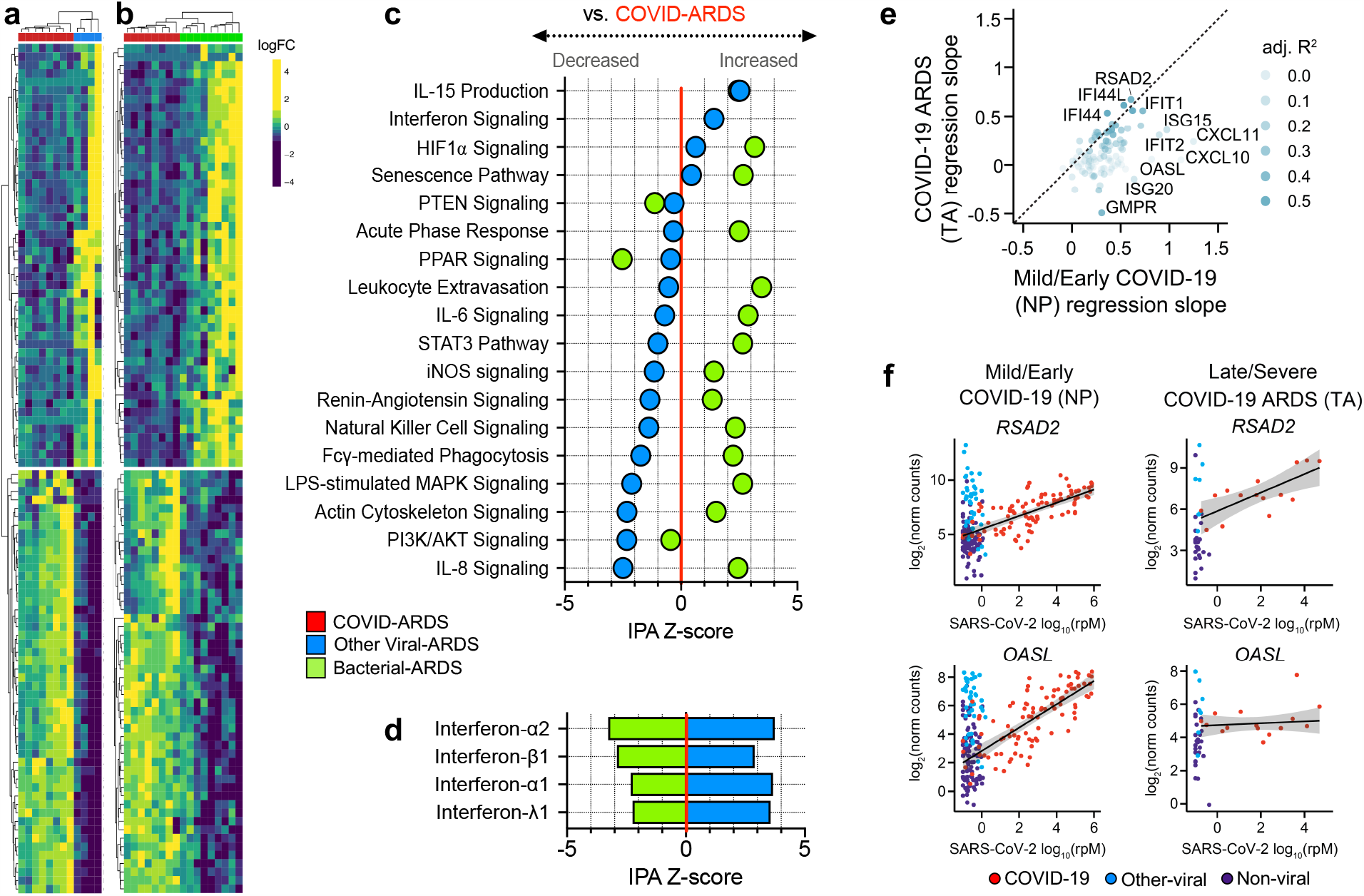
Lower respiratory tract transcriptional signature of ARDS due to COVID-19 versus other viral or bacterial lower respiratory tract infections. **a)** Heatmap depicting expression and unsupervised clustering of top differentially expressed genes by adjusted P value between patients with COVID-19 related ARDS (COVID-ARDS, red) versus ARDS due to viral LRTI (Viral-ARDS, blue). **b)** Heatmap depicting expression and unsupervised clustering of the top 50 differentially expressed genes between patients with COVID-19 related ARDS (COVID-ARDS, red) versus ARDS due to bacterial LRTI (Bacterial-ARDS, green). **c)** Pathway analysis based on differentially expressed genes depicting relative expression of signaling pathways by IPA Z-score with respect to a baseline of gene expression in COVID-ARDS. Values are tabulated in (Supplementary Data 8). **d)** Predicted activation of upstream interferons in patients with ARDS due to viral or bacterial LRTI compared to those with COVID-ARDS revealed downregulation of type-I/III interferons in COVID-ARDS versus other viral LRTI-related ARDS. Values tabulated in (Supplementary Data 9). **e)** Scatterplot of the relationship between interferon-stimulated gene (ISG) counts and SARS-CoV-2 viral load (reads per million, rpM), quantified by the regression slope, in nasopharyngeal (NP) samples from patients with mostly mild/early COVID-19 (x-axis) and in tracheal aspirate (TA) samples from patients with severe COVID-19 and ARDS (y-axis). **f)** *RSAD2* is an ISG whose expression (y-axis) is correlated with SARS-CoV-2 viral load (x-axis) in both early/mild (NP) and severe (TA) disease, while *OASL* is an ISG for which the correlation observed in early/mild COVID-19 is absent in severe COVID-19 patients with ARDS. Values are tabulated in (Supplementary Data 10).

Although interferon-related gene expression was higher in COVID-ARDS compared to bacterial LRTIs and no-ARDS controls, it was markedly attenuated in ARDS patients with COVID-19 versus those with other viral LRTI (**Figure 2d, Supplementary Data 10, Supplementary Methods**). Prior studies found strong correlation between SARS-CoV-2 viral load and expression of interferon-stimulated genes (ISGs) in the upper respiratory tract of patients with mild disease, early during infection^16^. In contrast, in the lower respiratory tract of patients with severe disease, we observed decoupling of this relationship for several ISGs (**Figures 2e-f, Supplementary Data 11**), suggesting that a dysregulated interferon response in the lower respiratory tract may characterize severe COVID-19. This hypothesis is supported by recent findings of impaired interferon signaling in peripheral blood immune cells of patients with severe versus mild COVID-19^17^, and a recent report suggesting that a dysregulated interferon response may be a universal characteristic of severe viral infections^18^.

## Discussion

Our results challenge the cytokine storm model of COVID-19 ARDS. Instead, we observe a complex picture of host immune dysregulation that includes upregulation of genes with non-canonical roles in inflammation, immunity and interferon signaling that are predicted to be attenuated by dexamethasone, G-CSF and other potential therapeutics. Our work emphasizes the value of including clinically relevant control patients in COVID-19 immunophenotyping studies and underscores that detection of elevated cytokine levels in the blood does not necessarily equate to causality in pathogenesis. Single cytokine blockade was attempted unsuccessfully in the past for treatment of sepsis^19^, which like COVID-19, is characterized by dysregulated host response to infection as well as significant biologically meaningful heterogeneity^20^.

This work builds on recent reports of dysregulated interferon responses in patients with severe COVID-19 pneumonia and suggests that decoupling of viral load from interferon signaling may be a relevant pathologic feature of severe disease. Further work in a larger cohort that also includes direct measurement of cytokine levels in the lower airway will be needed to validate these results. In conclusion, comparative lower respiratory transcriptional profiling of patients with COVID-19 and other ARDS etiologies did not find evidence of cytokine storm but did reveal potential therapeutic agents and novel gene targets that may mitigate the dysregulated host response of COVID-ARDS.

## Data and Code Availability

Human gene counts for the samples generated in this study can be obtained under NCBI GEO accession GSE163426. The published human lung single-cell datasets^17^ used for cell type proportions analysis can be obtained through Synapse under accessions syn21560510 and syn21560511. Code for the differential expression and cell type proportions analysis is available at: https://github.com/AartikSarma/COVIDARDS.

## Data Availability

Human gene counts for the samples generated in this study can be obtained under NCBI GEO accession GSE163426. The published human lung single-cell datasets used for cell type proportions analysis can be obtained through Synapse under accessions syn21560510 and syn21560511. Code for the differential expression and cell type proportions analysis is available on GitHub (see link).

https://github.com/AartikSarma/COVIDARDS

## Acknowledgements

This study was performed with support from the National Institute of Allergy and Infectious Diseases-sponsored Immunophenotyping Assessment in a COVID-19 Cohort (IMPACC) Network. We gratefully appreciate support from Amy Kistler, PhD, Jack Kamm, PhD, Saharai Caldera, BS, Maira Phelps BS and Norma Neff, PhD. We thank Mark and Carrie Casey, Julia and Kevin Hartz, Carl Kawaja and Wendy Holcombe, Eric Keisman and Linda Nevin, Martin and Leesa Romo, Three Sisters Foundation, Diana Wagner and Jerry Yang and Akiko Yamazaki for their support.

## Author Contributions

CRL, CSC, AS and SC conceived and designed the study. TD, RG, PHS and KMA oversaw or performed sample processing, library preparation and sequencing. CSC, CMH, KNK RG, AJ, JGW and ERS coordinated or contributed to clinical operations including patient enrollment. CD, AS, CRL, FM, TD and NS performed metadata collation or clinical chart review. AS, EM, AOP, CRL and SAC performed data analysis and interpretation. CSC, SAC, EM, DJE, JLD, KMA, MFK, PFW, MAM, BSZ, JGW, AL, AB, FM, PS and MS provided guidance, advice and comments on the study design and manuscript. CRL, AS and CSC wrote the manuscript with input from all authors.

## Supplementary Materials

### Supplementary Methods

#### Study design, clinical cohort and ethics statement

We conducted a case-control study of patients with ARDS due to COVID-19 (n = 15) versus two control groups of either patients with ARDS due to other causes (n = 32) or patients intubated for airway protection without evidence of pulmonary pathology on imaging (n = 5). We studied patients who were enrolled in either of two prospective cohort studies of critically ill patients at the University of California, San Francisco (UCSF) and Zuckerberg San Francisco General Hospital. Both studies were approved by the UCSF Institutional Review Board according under protocols 17-24056 and 20-30497, respectively, which granted a waiver of initial consent for tracheal aspirate and blood sampling. Informed consent was subsequently obtained from patients or their surrogates for continued study participation, as previously described^1^.

For this analysis, inclusion criteria were: 1) admission to the intensive care unit for mechanical ventilation for ARDS or airway protection, 2) age ≥ 18 years, 3) availability of TA with 10^6^ protein-coding reads collected within five days of intubation. Exclusion criteria were: 1) receipt of immunosuppressive medication or underlying immunocompromising condition prior to tracheal aspirate collection. Subject charts and chest x-rays were reviewed by at least two study authors (AS, PS, ES, FM, CD, MM, CL, CC) to confirm a diagnosis of ARDS using the Berlin Definition^2^. Lower respiratory tract infections were adjudicated by two study physicians using the United States Centers for Disease Control surveillance definition of pneumonia^3^. Of 72 patients initially screened, nine with ARDS due to COVID-19 and 10 with ARDS due to other causes were excluded because of treatment with immunosuppressant medications or because of an underlying immunocompromising condition (e.g., solid organ transplantation, bone marrow transplantation, human immunodeficiency virus infection).

#### Metagenomic sequencing

Following enrollment, TA was collected and mixed 1:1 with DNA/RNA shield (Zymo Research) to preserve nucleic acid. To evaluate host gene expression and detect the presence of SARS-CoV-2 and other viruses, metagenomic next generation sequencing (mNGS) of RNA was performed on TA specimens. Following RNA extraction (Zymo Pathogen Magbead Kit) and DNase treatment, human cytosolic and mitochondrial ribosomal RNA was depleted using FastSelect (Qiagen). To control for background contamination, we included negative controls (water and HeLa cell RNA) as well as positive controls (spike-in RNA standards from the External RNA Controls Consortium (ERCC))^4^. RNA was then fragmented and underwent library preparation using the NEBNext Ultra II RNAseq Kit (New England Biolabs). Libraries underwent 146 nucleotide paired-end Illumina sequencing on an Illumina Novaseq 6000 instrument.

#### Host differential expression and pathway analysis

Following demultiplexing, sequencing reads were pseudo-aligned with kallisto^5^ (v. 0.46.1; including bias correction) to an index consisting of all transcripts associated with human protein coding genes (ENSEMBL v. 99), cytosolic and mitochondrial ribosomal RNA sequences, and the sequences of ERCC RNA standards. Samples retained in the dataset had a total of at least 1,000,000 estimated counts associated with transcripts of protein coding genes, and the median across all samples was 7.3 million. Gene-level counts were generated from the transcript-level abundance estimates using the R package tximport^6^, with the scaledTPM method.

Differential expression analysis was performed using DESeq2^7^. We modeled the expression of individual genes using the design formula ∼ARDSEtiology. In our primary analysis, the ARDS etiology was categorized as COVID-ARDS, Other-ARDS, or No-ARDS. In our secondary analysis, the ARDS etiology was categorized as COVID-ARDS, Viral-ARDS, Bacterial-ARDS, or No-ARDS. COVID-ARDS patients with viral or bacterial co-infections were excluded from this secondary analysis. Significant genes were identified using an independent-hypothesis-weighted, Benjamini-Hochberg false discovery rate (FDR) less than 0.1^8,9^. Empirical Bayesian adaptive shrinkage estimators for log_2_-fold change were fit using *ashr*^10^. We generated heatmaps of the top 50 differentially expressed genes by absolute log_2_-fold change. For visualization, gene expression was normalized using the variance stabilizing transformation, centered, and z-scaled. Heatmaps were generated using the *pheatmap* package. Patients were clustered using Euclidean distance and genes were clustered using Manhattan distance. Differentially expressed genes (FDR < 0.1) were analyzed using Ingenuity Pathway Analysis (IPA, Qiagen)^11,12^.

#### Canonical pathway analysis and drug/cytokine upstream regulator analysis

To evaluate signaling pathways and upstream transcriptional regulators from gene expression data, we employed IPA. Specifically, genes were analyzed using Core, Canonical Pathway and Upstream Regulator Analysis on shrunken log2-fold change. IPA Upstream Regulator Analysis was employed to identify potential drug and cytokine regulators and predict their activation states based on expected effects between regulators and their known target genes or proteins annotated in the Ingenuity Knowledge Base (IKB)^12^. IPA calculates a Fisher’s exact p-value for overlap of differentially expressed genes with curated gene sets representing canonical biological pathways, or upstream regulators of gene expression, including cytokines and 12,981 drugs. In addition, IPA calculates a Z-score for the direction of gene expression for a pathway or regulator based on the observed gene expression in the dataset. The Z-score signifies whether expression changes for genes within pathways, or for known target genes of upstream regulators, are consistent with what is expected based on previously published analyses annotated in the IKB. Significant pathways and upstream regulators were defined as those with a Z-score absolute value greater than 2 and an overlap P value < 0.05.

#### *In silico* analysis of cell type proportions

Cell-type proportions were estimated from bulk host transcriptome data using the CIBERSORT X algorithm^13^. We used the Human Lung Cell Atlas dataset^14^ to derive the single cell signatures. The cell types estimated with this reference cover all expected cell types in the airway. The estimated proportions were compared between the three patient groups using a Mann-Whitney-Wilcoxon test (two-sided) with Bonferroni correction.

#### Quantification of SARS-CoV-2 viral load by mNGS

All samples were processed through a SARS-CoV-2 reference-based assembly pipeline that involved removing reads likely originating from the human genome or from other viral genomes annotated in RefSeq with Kraken2 (v. 2.0.8_beta), and then aligning the remaining reads to the SARS-CoV-2 reference genome MN908947.3 using minimap2 (v. 2.17). We calculated SARS-CoV-2 reads-per-million (rpM) by dividing the number of reads that aligned to the virus with mapq ≥ 20 by the total number of reads in the sample (excluding reads mapping to ERCC RNA standards).

#### Regression of ISG counts against viral load in TA and NP samples

We assembled a set of 100 interferon-stimulated genes based on the “Hallmark interferon alpha response” gene set in MSigDB^15^. We then performed robust regression of the quantile normalized gene counts (log_2_ scale), generated using the R package *limma*, against log_10_(rpM) of SARS-CoV-2. This was done in two separate datasets of COVID-19 patients: i) the tracheal aspirate (TA) samples from patients with COVID-19 ARDS reported in this study (n=15); and ii) the nasopharyngeal swab (NP) samples from patients with mostly early and mild disease that we previously reported (n=93)^16^. The analysis was performed using the R package robustbase (v. 0.93.6), which implements MM-type estimators for linear regression. Model predictions were generated using the R package ggeffects (v. 0.14.3) and used for display in the individual gene plots. Error bands represent normal distribution 95% confidence intervals around each prediction. Reported P-values for significance of the difference of the regression coefficient from 0 are based on a t-statistic and Benjamini-Hochberg adjusted. Reported R^2^ values represent the adjusted robust coefficient of determination.

## Data and Code Availability

Human gene counts for the samples generated in this study can be obtained under NCBI GEO accession GSE163426. The published human lung single-cell datasets^17^ used for cell type proportions analysis can be obtained through Synapse under accessions syn21560510 and syn21560511. Code for the differential expression analyses, cell type proportions analysis and gene expression classifiers is available at: https://github.com/AartikSarma/COVIDARDS.

**Supplementary Table 1.** Clinical and demographic characteristics of patients with ARDS due to COVID-19 (COVID-ARDS), control patients with ARDS due to other etiologies (Other-ARDS), and intubated control patients without ARDS (No-ARDS).

|  | **COVID-ARDS** | **Other-ARDS** | **P** | **No-ARDS** | **P** |
| --- | --- | --- | --- | --- | --- |
| **N** | 15 | 32 |  | 5 |  |
| **Age (median [IQR])** | 54.8<br>[42.5, 67.5] | 61.4<br>[47.3, 71.5] | 0.205 | 66.2<br>[62.0, 82.0] | 0.190 |
| **Male** | 9 (60.0) | 20 (62.5) | 1.000 | 2 (40.0) | 0.795 |
| **30-day mortality** | 3 (20.0) | 11 (34.4) | 0.508 | 2 (40.0) | 0.546 |
| **Race (%)** |  |  | <0.001 |  | 0.029 |
| African American | 0 (0.0) | 2 (6.2) |  | 0 (0.0) |  |
| Asian | 3 (20.0) | 4 (12.5) |  | 1 (20.0) |  |
| Caucasian | 1 (6.7) | 23 (71.9) |  | 3 (60.0) |  |
| Other | 11 (73.3) | 3 (9.4) |  | 1 (20.0) |  |
| **Hispanic ethnicity** | 8 (53.3) | 3 (9.4) | 0.003 | 0 (0.0) | 0.114 |
| **PaO<sub>2</sub>/FiO<sub>2</sub> (median [IQR])\*** | 74.0<br>[60.5, 115.0] | 96.0<br>[67.0, 148.0] | 0.114 | 296.0<br>[216.0, 366.5] | 0.003 |
| **ARDS etiology (%)** |  |  | 0.109 |  | <0.001 |
| Aspiration | 0 (0.0) | 5 (15.6) |  | 0 (0.0) |  |
| LRTI | 15 (100.0) | 20 (62.5) |  | 0 (0.0) |  |
| Sepsis | 0 (0.0) | 4 (12.5) |  | 0 (0.0) |  |
| Transfusion | 0 (0.0) | 2 (6.2) |  | 0 (0.0) |  |
| Unknown | 0 (0.0) | 1 (3.1) |  | 0 (0.0) |  |
| None | 0 (0.0) | 0 (0.0) |  | 5 (100.0) |  |
| **LRTI type (%)** |  |  | <0.001 |  | <0.001 |
| Bacterial | 0 (0.0) | 9 (28.1) |  | 0 (0.0) |  |
| Viral | 8 (60.0) | 4 (12.5) |  | 0 (0.0) |  |
| Viral + Bacterial | 4 (20.0) | 3 (9.4) |  | 0 (0.0) |  |
| Viral + Viral | 3 (20.0) | 0 (0.0) |  | 0 (0.0) |  |
| Unknown | 0 (0.0) | 4 (12.5) |  | 0 (0.0) |  |
| None | 0 (0.0) | 12 (37.5) |  | 5 (100.0) |  |

P-values represent comparisons versus COVID-ARDS. Reasons for intubation of No-ARDS patients included: hemorrhagic stroke, subdural hematoma, retroperitoneal hemorrhage or other neurosurgical procedure. Statistical significance was determined using Fisher's exact test (discrete variables) or by Wilcoxon test (continuous variables). \*Lowest PaO<sub>2</sub>/FiO<sub>2</sub> recorded in first five days of mechanical ventilation. PF ratios were not available for two Other-ARDS subjects, who were diagnosed with ARDS based on an SaO<sub>2</sub>/FiO<sub>2</sub> < 315. IQR = Interquartile Range.

**Supplementary Figure 1.**
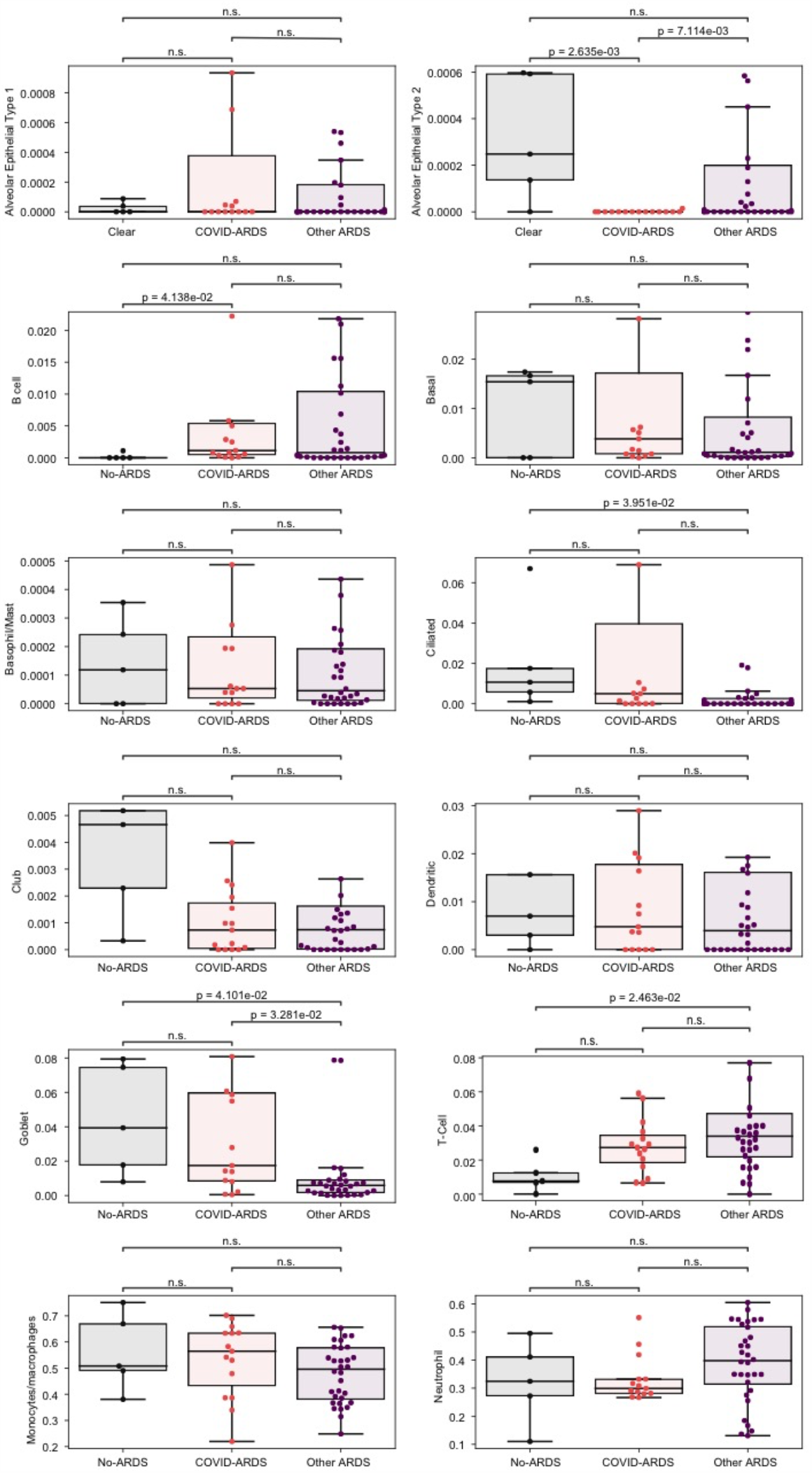
*In silico* deconvolution of cell types from tracheal aspirate bulk RNA-sequencing data using lung single cell signatures. The horizontal line inside the box denotes the median and the lower and upper hinges correspond to the first and third quartiles, respectively. Whiskers extend from the hinge to the largest (smallest, respectively) value no more than 1.5*IQR away from the hinge, where IQR is the interquartile range. The y-axis in each panel was trimmed at the maximum value among the three patient groups of 1.5*IQR above the third quartile. Pairwise comparisons between patient groups were performed with a two-sided Mann-Whitney-Wilcoxon test followed by Bonferroni’s correction (n=15 COVID-ARDS, n=32 Other ARDS, n=5 No-ARDS). Data are tabulated in (Supplementary Data 5).

**Supplementary Data 1**. Detailed clinical, microbiological and demographic features of the cohort. Legend: *SaO_2_/FiO_2_ Ratio (SF Ratio) < 315 was used to verify ARDS diagnosis in these subjects.

**Supplementary Data 2**. Differentially expressed genes (adjusted P value (padj) < 0.1) between patients with ARDS due to COVID-19 (COVID-ARDS) versus **a)** controls with ARDS due to other etiologies (Other-ARDS) or **b)** intubated controls without ARDS (No-ARDS). Positive fold change indicates gene is upregulated in COVID-ARDS.

**Supplementary Data 3**. Ingenuity Pathway Analysis (IPA) of differentially expressed genes (padj < 0.1) between patients with ARDS due to COVID-19 (COVID-ARDS) versus **a)** controls with ARDS due to other etiologies (Other-ARDS) or **b)** intubated controls without ARDS (No-ARDS). Positive Z-score indicates pathway is upregulated in COVID-ARDS. Pathways with a Z-score absolute value ≥ 1 are included in table.

**Supplementary Data 4**. Plasma concentrations of inflammatory cytokines in patients with ARDS due to COVID-19 (COVID-ARDS) versus controls with ARDS due to other etiologies (Other-ARDS).

**Supplementary Data 5**. *In silico* deconvolution of cell type proportions from tracheal aspirate bulk RNA-sequencing data using lung single cell signatures. Data are plotted in (Supplementary Figure 1).

**Supplementary Data 6**. Chemical and biological drugs computationally predicted by IPA to attenuate the transcriptional response of ARDS due to COVID-19 (COVID-ARDS) against a comparator group of **a)** ARDS due to other causes (Other-ARDS) or **b)** intubated controls without ARDS (No-ARDS). Drugs with a Z-score > 2 are included in table.

**Supplementary Data 7**. Genes affected by drugs computationally predicted by IPA to modulate the transcriptional response of ARDS due to COVID-19 (COVID-ARDS) against a comparator group of **a)** ARDS due to other causes (Other-ARDS) or **b)** intubated controls without ARDS (No-ARDS). Predicted transcriptional effect of each drug on each gene is indicated with respect to comparator group 1 in the table.

**Supplementary Data 8**. Differentially expressed genes (padj < 0.1) between patients with ARDS due to COVID-19 (COVID-ARDS) versus controls with ARDS due to **a)** other viral lower respiratory tract infections (Other Viral-ARDS) or **b)** bacterial lower respiratory tract infections (Bacterial-ARDS). Positive log_2_ fold change indicates gene is upregulated in COVID-ARDS.

**Supplementary Data 9**. Pathway analysis (IPA) of differentially expressed genes (padj < 0.1) between patients with ARDS due to COVID-19 (COVID-ARDS) versus controls with ARDS due to other viral or bacterial lower respiratory tract infections. Z-scores are with respect to COVID-ARDS. Pathways with a Z-score absolute value ≥ 1 are included in table.

**Supplementary Data 10**. Computationally predicted (IPA) upstream cytokines based on the transcriptional signature of COVID-19 ARDS (COVID-ARDS) compared to ARDS from other viral LRTI. Pathways with a Z-score absolute value ≥ 1 are included in table.

**Supplementary Data 11**. Relationship between interferon-stimulated gene (ISG) expression and SARS-CoV-2 viral load measured in RNA-seq reads per million (rpM) for COVID-19 patients with late, severe disease and ARDS (COVID-ARDS) from lower respiratory tract samples (this study), and COVID-19 patients with early, mostly mild disease measured from upper respiratory tract samples^16^. Legend: reg_intercept = intercept in the robust regression of gene expression on SARS-CoV-2 viral load; reg_slope = slope in the robust regression of gene expression on SARS-CoV-2 viral load; reg_adj_R2 = adjusted robust coefficient of determination; reg_p_adj = Benjamini-Hochberg adjusted p-value for difference of the regression slope from 0.

**Supplementary Appendix**. COMET Consortium Member list.

## References

1. Matthay, M. A. et al. Acute respiratory distress syndrome. Nature Reviews Disease Primers 5, (2019).

2. Mehta, P. et al. COVID-19: consider cytokine storm syndromes and immunosuppression. The Lancet 395, 1033–1034 (2020).

3. Wilson, J. G. et al. Cytokine profile in plasma of severe COVID-19 does not differ from ARDS and sepsis. JCI Insight 5, (2020).

4. The RECOVERY Collaborative Group. Dexamethasone in Hospitalized Patients with Covid-19 — Preliminary Report. New England Journal of Medicine (2020) doi:10.1056/NEJMoa2021436.

5. Bost, P. et al. Host-Viral Infection Maps Reveal Signatures of Severe COVID-19 Patients. Cell 181, 1475-1488.e12 (2020).

6. Chua, R. L. et al. COVID-19 severity correlates with airway epithelium–immune cell interactions identified by single-cell analysis. Nature Biotechnology 38, 970–979 (2020).

7. Krämer, A., Green, J., Pollard, J. & Tugendreich, S. Causal analysis approaches in Ingenuity Pathway Analysis. Bioinformatics 30, 523–530 (2014).

8. Sesma, J. I. et al. UDP-glucose promotes neutrophil recruitment in the lung. Purinergic Signalling 12, 627–635 (2016).

9. Cai, W. et al. STAT6/Arg1 promotes microglia/macrophage efferocytosis and inflammation resolution in stroke mice. JCI Insight 4, (2019).

10. Chen, L. & Guo, D. The functions of tumor suppressor PTEN in innate and adaptive immunity. Cellular & Molecular Immunology 14, 581–589 (2017).

11. Schabbauer, G. et al. Myeloid PTEN Promotes Inflammation but Impairs Bactericidal Activities during Murine Pneumococcal Pneumonia. The Journal of Immunology 185, 468–476 (2010).

12. HCA Lung Biological Network et al. SARS-CoV-2 entry factors are highly expressed in nasal epithelial cells together with innate immune genes. Nature Medicine 26, 681–687 (2020).

13. Cheng, L. et al. Effect of Recombinant Human Granulocyte Colony–Stimulating Factor for Patients With Coronavirus Disease 2019 (COVID-19) and Lymphopenia: A Randomized Clinical Trial. JAMA Internal Medicine (2020) doi:10.1001/jamainternmed.2020.5503.

14. Kim, Y.-H., Lee, J.-R. & Hahn, M.-J. Regulation of inflammatory gene expression in macrophages by epithelial-stromal interaction 1 (Epsti1). Biochem Biophys Res Commun 496, 778–783 (2018).

15. Kaplan, M. H. STAT signaling in inflammation. JAK-STAT 2, e24198 (2013).

16. Mick, E. et al. Upper airway gene expression reveals suppressed immune responses to SARS-CoV-2 compared with other respiratory viruses. Nature Communications 11, (2020).

17. Hadjadj, J. et al. Impaired type I interferon activity and inflammatory responses in severe COVID-19 patients. Science 369, 718–724 (2020).

18. Zheng, H. et al. Multi-cohort analysis of host immune response identifies conserved protective and detrimental modules associated with severity irrespective of virus. http://medrxiv.org/lookup/doi/10.1101/2020.10.02.20205880 (2020) doi:10.1101/2020.10.02.20205880.

19. Cavaillon, J., Singer, M. & Skirecki, T. Sepsis therapies: learning from 30 years of failure of translational research to propose new leads. EMBO Molecular Medicine 12, (2020).

20. Seymour, C. W. et al. Assessment of Clinical Criteria for Sepsis: For the Third International Consensus Definitions for Sepsis and Septic Shock (Sepsis-3). JAMA 315, 762 (2016).

## Supplementary References

1. Langelier, C. et al. Integrating host response and unbiased microbe detection for lower respiratory tract infection diagnosis in critically ill adults. Proc Natl Acad Sci USA 201809700 (2018) doi:10.1073/pnas.1809700115.

2. ARDS Definition Task Force. Acute respiratory distress syndrome: the Berlin definition. JAMA 307, 2526–2533 (2012).

3. United States Centers for Disease Control and Prevention. CDC/NHSN Surveillance Definitions for Specific Types of Infections. (2017).

4. Pine, P. S. et al. Evaluation of the External RNA Controls Consortium (ERCC) reference material using a modified Latin square design. BMC Biotechnology 16, 54 (2016).

5. Bray, N. L., Pimentel, H., Melsted, P. & Pachter, L. Near-optimal probabilistic RNA-seq quantification. Nature Biotechnology 34, 525 (2016).

6. Soneson, C., Love, M. I. & Robinson, M. D. Differential analyses for RNA-seq: transcript-level estimates improve gene-level inferences. F1000Research 4, 1521 (2015).

7. Love, M. I., Huber, W. & Anders, S. Moderated estimation of fold change and dispersion for RNA-seq data with DESeq2. Genome Biology 15, 550 (2014).

8. Ignatiadis, N., Klaus, B., Zaugg, J. B. & Huber, W. Data-driven hypothesis weighting increases detection power in genome-scale multiple testing. Nature Methods 13, 577–580 (2016).

9. Benjamini, Y. & Hochberg, Y. Controlling the False Discovery Rate: A Practical and Powerful Approach to Multiple Testing. Journal of the Royal Statistical Society: Series B (Methodological) 57, 289–300 (1995).

10. Stephens, M. False discovery rates: a new deal. Biostatistics 18, 275–294 (2017).

11. Krämer, A., Green, J., Pollard, J. & Tugendreich, S. Causal analysis approaches in Ingenuity Pathway Analysis. Bioinformatics 30, 523–530 (2014).

12. Ingenuity Systems. Ingenuity Upstream Regulator Analysis in IPA. http://pages.ingenuity.com/rs/ingenuity/images/0812%20upstream_regulator_analysis_whitepaper.pdf.

13. Newman, A. M. et al. Robust enumeration of cell subsets from tissue expression profiles. Nature methods 12, 453–457 (2015).

14. Travaglini, K. J. et al. A molecular cell atlas of the human lung from single-cell RNA sequencing. Nature 587, 619–625 (2020).

15. Subramanian, A. et al. Gene set enrichment analysis: A knowledge-based approach for interpreting genome-wide expression profiles. Proceedings of the National Academy of Sciences 102, 15545–15550 (2005).

17. Travaglini, K. J. et al. A molecular cell atlas of the human lung from single cell RNA sequencing. bioRxiv 742320 (2020) doi:10.1101/742320.

